# Peripheral Neuropathy in Cancer Patients- Multifactorial Contributors to Dose Limiting and Chronic Toxicity

**DOI:** 10.1101/2024.07.24.24310956

**Authors:** Tiffany Li, Hannah C Timmins, Lisa G Horvath, Michelle Harrison, Peter Grimison, Michael Friedlander, Gavin Marx, Frances Boyle, David Wyld, Robert Henderson, Tracy King, Sally Baron-Hay, Matthew C Kiernan, Elizabeth H Barnes, David Goldstein, Susanna B Park

## Abstract

**Background and Objective:** Chemotherapy-induced peripheral neuropathy (CIPN) is a complex and dose-limiting toxicity of anticancer treatments with chronic symptoms leading to increased disability and reduced quality of life. The present study evaluated clinical risk factors associated with development of chronic, severe and dose-limiting CIPN, utilising a comprehensive multi-modal battery of neuropathy assessment.

**Methods:** Baseline clinical risk factors were investigated in patients who had completed neurotoxic chemotherapy (including taxanes, platinums and haematological cancer therapies). CIPN was assessed using neurological evaluation (Total Neuropathy Score, sural nerve conduction studies), patient reported outcome measure (EORTC QLQ-CIPN20), and clinically graded neuropathy (NCI-CTCAE). Multivariate models of risk factors associated with development of chronic, severe and dose-limiting CIPN were evaluated using backwards stepwise regression model building.

**Results:** The study recruited 903 patients (age 61 (IQR 50-69) years) who were assessed 12 (IQR 6-24) months post neurotoxic treatment. 73% of patients presented with CIPN at time of assessment, with 37% having moderate to severe symptoms. 32% of patients experienced neurotoxic treatment dose modification due to CIPN. Across the various CIPN assessment approaches, risk factors for chronic CIPN included older age, diabetes diagnosis, higher BMI and prior exposure to neurotoxic treatment (all P<0.05). Risk factors for severe CIPN included older age, higher BMI, prior neurotoxic treatment and female sex (all P<0.05), whereas risk factors for dose-limiting CIPN included older age and female sex (all P<0.05).

**Discussion:** This study identified baseline clinical risk factors associated chronic, severe and dose-limiting CIPN. Closer monitoring of these vulnerable cohorts will allow for timely CIPN management, including referral pathways to intervention and rehabilitation therapies which will ultimately lead to improved CIPN morbidity.

## Introduction

Neurological consequences of anticancer treatment have a significant impact on the growing global population of cancer survivors (1). Complex adverse events such as chemotherapy-induced peripheral neurotoxicity (CIPN) affect people treated for common cancers including colorectal, breast, gynaecological and blood cancers (2). Cardinal symptoms include numbness, tingling and shooting or burning pain at the distal extremities which may progress proximally (3), while motor and autonomic neuropathy occur less commonly (3). During treatment, development of CIPN often necessitates dose reduction or early treatment cessation (4), resulting in reduced exposure to anticancer treatment. Chronically, CIPN produces persistent sensory neuropathy and leads to increased falls risk, prolonged sleep disturbance and increased opioid use, all of which significantly and negatively impact on patients’ quality of life (QoL) (5).

Unfortunately, CIPN is highly prevalent, with >80% patients developing neurological signs and symptoms during neurotoxic treatment (6, 7) and up to 30-40% of patients requiring dose modification due to neuropathy (6, 8). While some patients demonstrate recovery, CIPN persists in up to 50% long term (6, 9). Ultimately, CIPN produces peripheral axon degeneration but specific mechanisms differ between agents (10) and the extent of degeneration may vary depending on individual metabolic and demographic features. Given the impact of CIPN on anticancer treatment and long-term QoL, it is critical to understand which patients are most at risk of neurological sequelae. However, inconsistent use of neuropathy outcome measures have limited our ability to identify robust clinical risk factors for CIPN across settings (11–14). Accordingly, it is necessary to interrogate CIPN risk factors in large-scale clinical cohorts, using comprehensive and multi-modal CIPN assessment tools.

The aim of this study was to evaluate the association between clinical factors that are routinely available at the commencement of cancer therapy, and the development of chronic, severe and significant neuropathy during treatment by utilising multiple approaches to CIPN assessment.

## Methods

### Study Design and Participants

Adult cancer patients who completed a comprehensive neuropathy assessment following completion of a neurotoxic chemotherapy regimen were eligible to participate in the study. Patients were recruited by their clinical care team in oncology centres in Sydney and Brisbane, Australia from July 2015 to December 2021 and were included into the study if they were treated with neurotoxic anticancer treatments (including taxanes, platinums, vinca-alkaloids, bortezomib and thalidomide). Patients were excluded from analysis if they did not receive at least two doses of neurotoxic treatment. Written informed consent was obtained in accordance with the Declaration of Helsinki, with studies approved by the Sydney Local Health District and South-Eastern Sydney Local Health District Human Research Ethics Committees.

### Neuropathy Assessment

A multi-modal battery of assessments was utilised to assess CIPN, consisting of a patient reported outcome measure, neurological assessment and clinical grading.

#### Patient reported CIPN

The validated patient reported CIPN measure, The European Organization for Research and Treatment of Cancer Quality of Life Chemotherapy-Induced Peripheral Neuropathy Questionnaire (EORTC QLQ-CIPN20) (15, 16) was used to evaluate CIPN from the patient perspective. This measure consists of 20 items, each scored on a 4-point scale (1- Not at all; 2- A little bit; 3- Quite a bit; 4- Very much). Final scores were linearly converted to a 0-100 scale, with higher scores indicating worse CIPN.

#### Neurologically evaluated CIPN

The Total Neuropathy Score, clinical version (TNSc, Johns Hopkins University ©) (17) is a validated composite validated measure of CIPN (18, 19) consisting of two patient reported items assessing the extent of sensory and motor symptoms, followed by four items assessing neuropathic signs including pin-prick and vibration sense, strength and tendon reflexes. Pinprick examinations were completed using Neurotips (Owens Mumford, Woodstock, UK), and vibration sensation was assessed using a Rydel-Seiffer tuning fork. Each item was scored 0-4 and summed for a total of score range of 0-24, where higher scores indicate worse neuropathy.

Nerve conduction studies were completed on the left sural nerve using conventional techniques (20) and a Nicolet EDX Synergy device (Natus Medical, Inc., Pleasanton, California). Antidromic sural sensory nerve action potentials (SNAPs) were recorded at the lateral malleolus with the stimulation site 10–15 cm proximal. Nerve amplitudes were compared to the lower-limit of age matched normative values (20).

#### Clinical grading of CIPN

was evaluated by trained researchers using the National Cancer Institute Common Terminology Criteria for Adverse Events (NCI-CTCAE) sensory neuropathy subscale version 3 (21). This measure discretely categorises CIPN severity on a scale of 0 to 4 (0- no symptoms; 1- asymptomatic, not interfering with daily function; 2- moderate symptoms, limiting daily function; 3- severe symptoms, limiting daily function and self-care; 4- disabling).

### Clinical Risk Factors

Demographic and clinical information including age, sex, body mass index (BMI; prior to commencing neurotoxic treatment), diabetic status, pre-existing neuropathy, prior exposure to neurotoxic treatments, cancer type and treatment information, were retrieved from medical records. Patient’s age was further categorised into <60, or ≥60 years and BMI was categorised as underweight/normal (BMI <25.0), overweight (BMI 25-29.9) or obese (BMI ≥30). Reasons for neurotoxic treatment dose modification were recorded with patients categorised as having ‘dose modification due to neuropathy’ if CIPN was the reason for dose reduction or early treatment cessation. Patients were classified into neurotoxic treatment groups including taxanes (paclitaxel, docetaxel, abraxane), platinum (oxaliplatin, cisplatin) or haematological cancer therapies (bortezomib, thalidomide, vinca-alkaloids) according to the highest cumulative dose of neurotoxic treatment received.

### Definitions

Risk factors were investigated for CIPN according to the following definitions. Dose-limiting CIPN was used as a surrogate for significant neuropathy development during treatment and defined as patients who received any dose modification or premature cessation of their neurotoxic treatment due to CIPN.

Chronic CIPN was defined in different ways depending on the assessment tool. This includes the presence of any grade on the NCI-CTCAE (NCI-CTCAE>0) or a reduction of sural SNAP amplitudes compared to normative controls (20) on nerve conduction studies. For the continuous CIPN outcome measures, chronic CIPN was assessed via higher scores on the TNSc and EORTC-CIPN20.

Severe CIPN was also defined in different ways depending on the assessment tool, including the highest quartile scores on the EORTC-CIPN20 and TNSc, and moderate-severe clinically graded CIPN (NCI-CTCAE ≥2).

### Statistical Analysis

All statistical analyses were conducted using Stata version 14 (StataCorp, College Station, Texas, USA). Descriptive data is presented as means with standard deviations or median (interquartile range) for parametric and non-parametric data, determined by the Shapiro-Wilk test of normality. Analysis of variance was used to compare CIPN outcome measures across patients with none (NCI-CTCAE=0), mild (NCI-CTCAE=1) and moderate/severe (NCI-CTCAE≥2) neuropathy. Significance was set to P<0.05.

Correlations between outcome measures were calculated using Spearman correlation, with r>0.6 indicating moderate correlations. Agreement between severe neuropathy for each outcome measure (NCI≥2, highest quartiles of EORTC-CIPN20 and TNSc) was investigated using the Kappa statistic, with κ<0.6 indicating weak agreement κ≥0.6 indicating moderate agreement (22).

Logistic regression (for binary outcomes) and linear regression (for continuous outcomes) adjusting for time since end of neurotoxic chemotherapy were conducted to investigate individual of clinical risk factors associated with CIPN. Multivariate logistic and linear regression models were fitted with backwards stepwise variable selection to investigate the overall association between risk factors and CIPN. The initial model included all clinical risk factors. Candidate risk factors were subsequently removed one-by-one starting with the least significant, until only predictors with P-value less than 0.05 remained in the model (23). Odds ratio (OR) for dichotomised outcomes (NCI-CTCAE, abnormal sural, dose modification due to CIPN) and β coefficient for continuous outcomes (TNSc, EORTC-CIPN20) and associated 95% confidence intervals were computed for all models.

## Results

A total of 901 patients were assessed following neurotoxic cancer therapy, at a median of 12 (6–24) months post completion. Patients had a median age of 61 (50–69) years, with 52% (n=468) aged ≥60 years. The majority of patients were female (66%, n=597) with breast cancer (33%, n=299). Taxanes were the most commonly received chemotherapies (57%, n=518), followed by platinum (32%, n=285) and haematological cancer therapies (11%, n=98). Further patient demographic information is detailed in Table 1.

**Table 1.** Demographic and clinical information.

| **N=901** |  |
| --- | --- |
|  | **Median (IQR)** |
| Age (years) | 61 (19) |
| Time since treatment (months) | 12 (18) |
|  | **N (%)** |
| Gender (female) | 596 (66%) |
| BMI (kg/m<sup>2</sup>) |  |
| Underweight/Healthy (<25) | 331 (38%) |
| Overweight (BMI 25 - 29.9) | 328 (38%) |
| Obese (BMI ≥30) | 207 (24%) |
| Not recorded | 35 (4%) |
| Diabetes (Yes) | 83 (9%) |
| Prior neurotoxic chemotherapy (Yes) | 57 (6%) |
| Pre-Existing Neuropathy (Yes) | 17 (2%) |
| Dose modification due to neuropathy (Yes)<sup>1</sup> | 275 (31%) |
| **Cancer Type** |  |
| Breast | 299 (33%) |
| Gastrointestinal | 235 (26%) |
| Gynaecological | 151 (17%) |
| Haematological | 94 (10%) |
| Prostate | 39 (4%) |
| Testicular | 30 (3%) |
| Head and Neck | 19 (2%) |
| Other<sup>2</sup> | 34 (4%) |
| **Cancer Stage** |  |
| 0-III | 594 (66%) |
| IV | 184 (20%) |
| Non-solid tumour | 94 (10%) |
| Unknown | 29 (3%) |
| **Neurotoxic Chemotherapy Type** |  |
| Taxanes | 518 (57%) |
| Platinum | 285 (32%) |
| Haematological chemotherapies | 98 (11%) |

<sup>1</sup>Dose modification information available for 879 patients; <sup>2</sup>Other cancer types include lung, peritoneum, bone, oesophagus, brain, liver, unknown

### Neuropathy burden

Dose modification information (including reduction or early cessation) for neurotoxic treatment was available for 879 patients, 31% (n=275) of which received dose modification due CIPN, i.e. developed dose limiting CIPN. There were no differences between the rates of dose modification due to CIPN between the chemotherapy types (Taxanes 33% n=169/509, Platinum 31% n=87/279, Haematological cancer therapies 21% n=19/91; χ²=5.5, P>0.05)

Majority of patients presented with CIPN (72.6%, 654/901), with 35.2% (n=317) having mild (grade 1) and 37.4% (n=337) having moderate to severe CIPN (grade ≥2) graded on the NCI-CTCAE. Of the patients with CIPN, 62.2% (407/654) reported functional impacts resulting from CIPN symptoms (including difficulties with feeling ground under feet, holding pen, manipulating small objects in hands). Increased time since treatment completion at time of assessment was associated with reduced patient reported, neurologically and clinically evaluated CIPN (P<0.05, Supplementary Table 1). There was also an association between chemotherapy type and CIPN severity, with platinum-treated patients overall presenting with worse chronic symptoms (all P<0.05, Supplementary Table 2). Patient reported and neurological evaluated CIPN indicated worse neuropathy outcomes for patients with higher clinically graded CIPN (P<0.05; Figure 1). Nerve conduction studies were completed on 743 patients, with 34.9% (n=259) presenting with reduced sural SNAPs compared to age-matched normative values (normal vs reduced median sural SNAP amplitudes 11.4 (8.5-16.8) µV vs 3.5 (1.1 – 4.6) µV). Patients presenting with neuropathy (NCI-CTCAE>0) were more likely to have reduced sural SNAPs (χ²=44.1, P<0.001).

**Figure 1.**
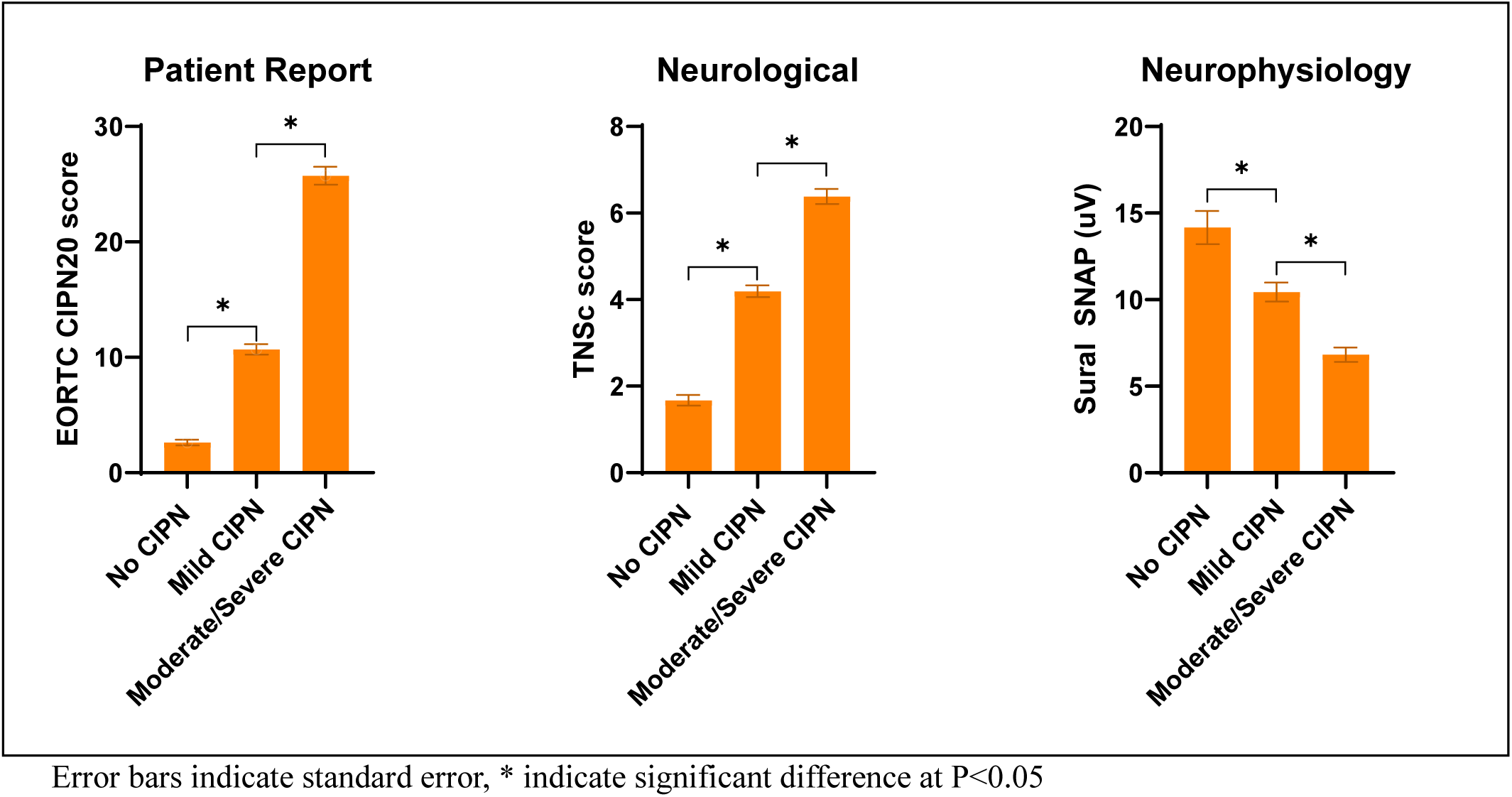
Patient reported, neurological and neurophysiological assessment of CIPN by clinical neuropathy grading. No CIPN NCI-CTCAE=0, Mild CIPN NCI-CTCAE=1, Moderate/severe CIPN NCI-CTCAE≥2.

### Neuropathy assessment agreement

Clinical, patient reported and neurologically assessed neuropathy overall reflected similar levels of chronic CIPN, with moderate correlation between these outcomes (NCI-CTCAE, EORTC-CIPN20, TNSc all r>0.6, P<0.001; Figure 2). However, this agreement delineates in their categorisation of severe CIPN (weak agreement between highest quartile of EORTC-CIPN20 and TNSc, NCI-CTCAE≥2, κ= 0.36 – 0.56; Figure 3) suggesting each assessment approach captures severe CIPN differently.

**Figure 2.**
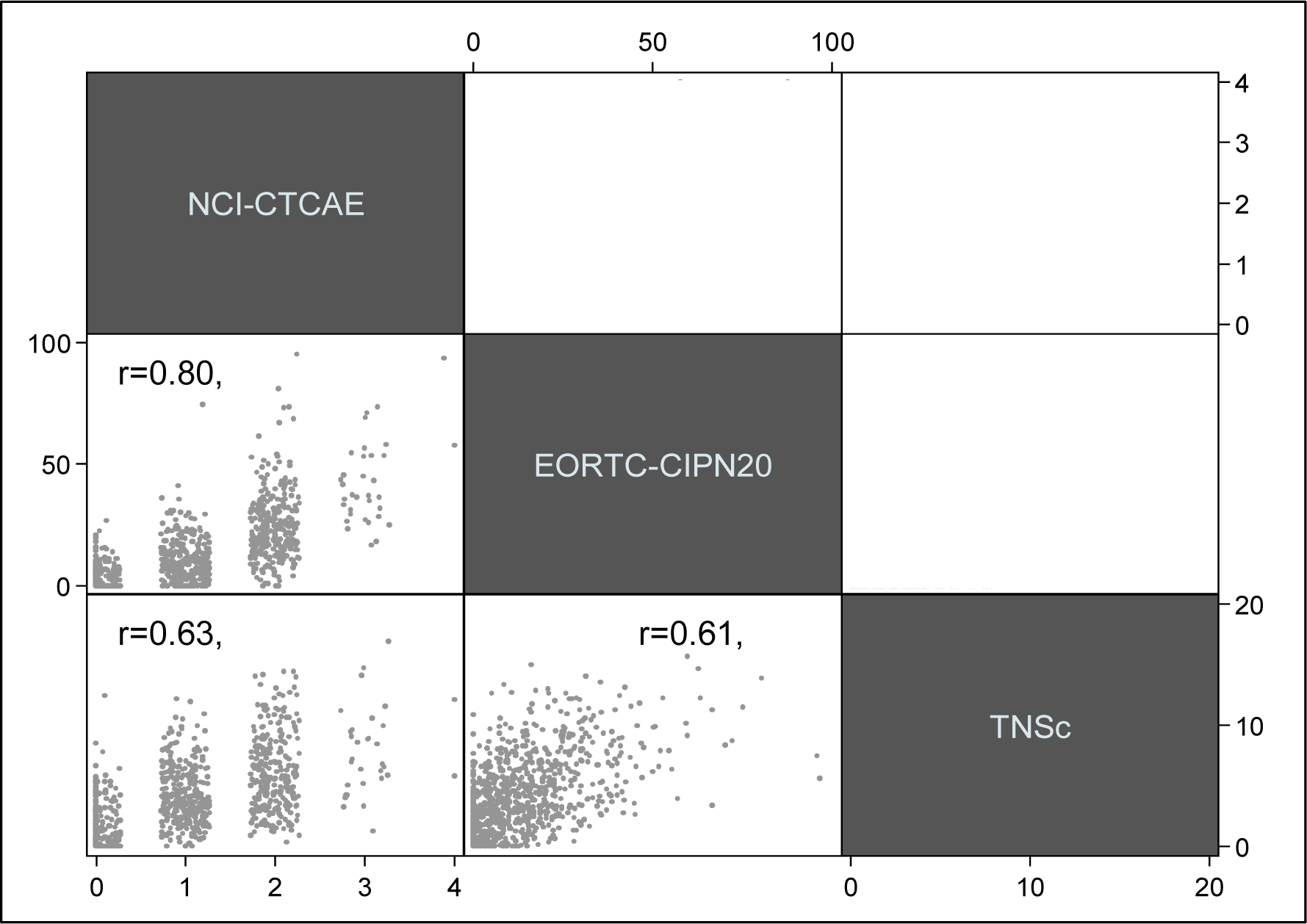
Scatterplot matrix between CIPN outcome measure scores.

**Figure 3.**
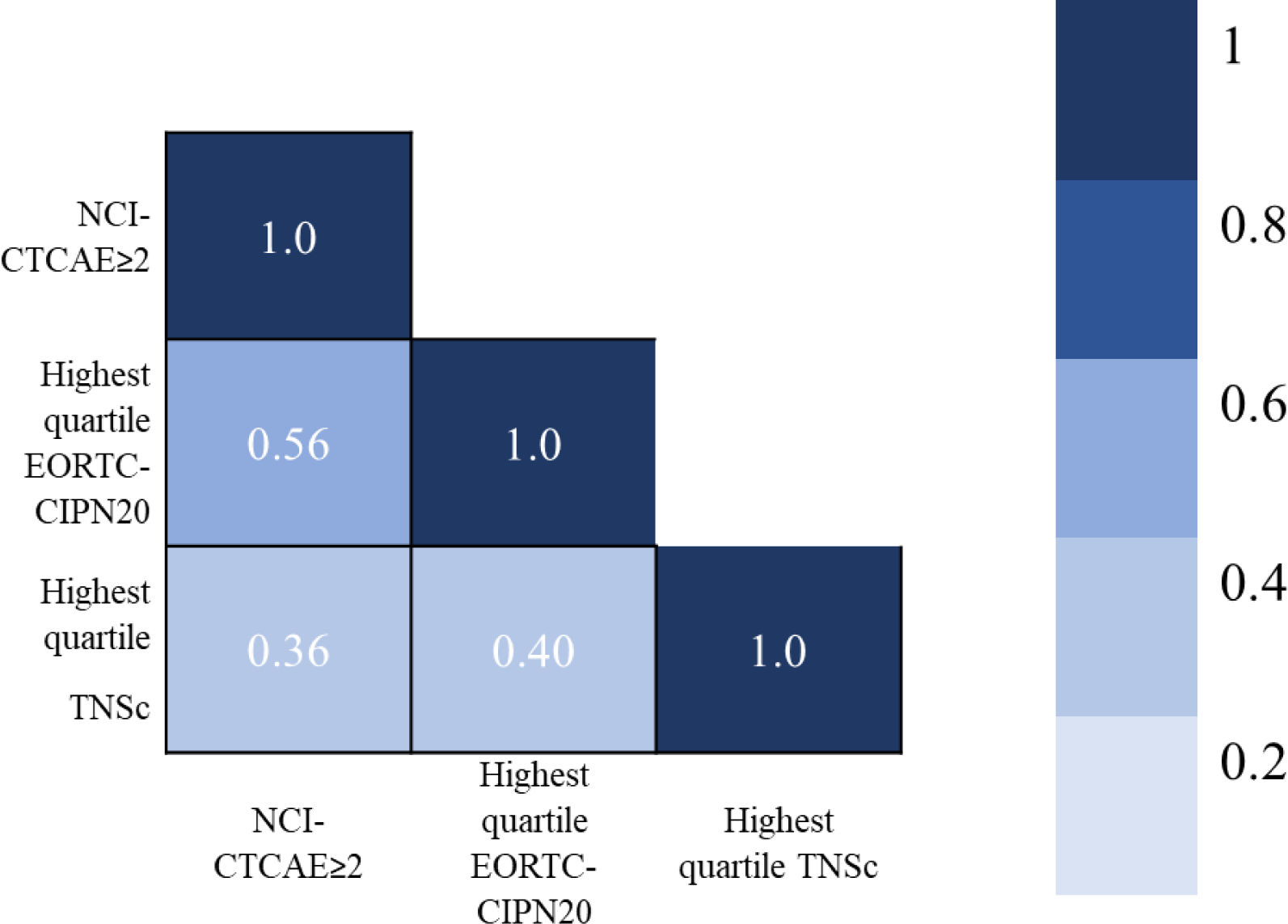
Matrix indicating agreement between high-grade neuropathy assessing using different CIPN outcome measures, presented as Kappa’s statistic. P<0.001 for all values.

### Risk factors for dose-limiting CIPN

Dose modification of neurotoxic treatment due to CIPN was used as a surrogate to investigate risk factors associated with development of significant CIPN during treatment. Information regarding dose modification was available for 879 patients, of which 31.3% (n=275) received neurotoxic treatment dose modification due to CIPN.

On univariate logistic regression (Table 2), female sex was the only significant risk factor for dose-limiting CIPN (OR=1.7 95% CI 1.3 – 2.4). Backwards stepwise multivariate regression models were subsequently built, initially including all clinical risk factors. Candidate risk factors were then removed one-by-one starting with the least significant, until only significant predictors remained in the model. After controlling for chemotherapy type, multivariate analysis indicated older age (≥60 years, OR=1.4 95% CI 1.0 – 1.8) and female sex (OR=1.9 95% CI 1.3 – 2.8) as significant risk factors for dose-limiting CIPN (Table 2).

**Table 2.** Univariate and multivariate models of risk factors for developing significant acute CIPN, using dose modification of neurotoxic treatment due to CIPN as a surrogate outcome measure^1^.

| Risk Factor | Dose Modification due to CIPN |  |  |  |
| --- | --- | --- | --- | --- |
|  | Univariate analysis |  | Multivariate analysis<sup>2</sup> |  |
|  | Odds ratio<br>(95% CI) | P | Odds ratio<br>(95% CI) | P |
| Age ($\geq 60$ vs $< 60$ years) | 1.3 (1.0 – 1.7) | $> 0.05$ | 1.4 (1.0 – 1.8) | $< 0.05^*$ |
| Diabetes (Yes vs No) | 1.5 (0.9 – 2.4) | $> 0.05$ | - | - |
| BMI (compared to<br>underweight/normal) |  |  |  |  |
| Overweight | 1.1 (0.8 – 1.5) | $> 0.05$ | - | - |
| Obese | 1.1 (0.8 – 1.6) | $> 0.05$ | - | - |
| Prior neurotoxic treatment<br>(Yes vs No) | 1.0 (0.6 – 1.8) | $> 0.05$ | - | - |
| Sex (female vs male) | 1.7 (1.3 – 2.4) | $< 0.005^*$ | 1.9 (1.3 – 2.8) | $< 0.005^*$ |
| Pre-existing neuropathy (Yes<br>vs No) | 1.2 (0.4 – 3.3) | $> 0.05$ | - | - |

\*Indicates significance at $P < 0.05$ .
<sup>1</sup>Dose modification information available for 879 participants, of which 275 (31.3%) received dose modification due to CIPN
<sup>2</sup>Multivariate model controls for chemotherapy type

### Risk factors for chronic CIPN

Linear and logistic regressions to investigate individual risk factors for chronic CIPN assessed using EORTC-CIPN20, TNSc, NCS and NCI-CTCAE, were computed, controlling for time since treatment completion (Supplementary Table 3). Older age (≥60 years), diabetes and higher BMI (overweight and obese compared to underweight/ healthy) were significant risk factors across all four measures of CIPN (P<0.05).

Multivariate regression analyses were subsequently investigated to identify risk factors associated with chronic CIPN (assessed via EORTC-CIPN20, TNSc, NCS and NCI-CTCAE), controlling for time since neurotoxic treatment and chemotherapy type. Final models only including significant risk factors (P<0.05) are presented in Table 3. Overall, significant risk factors for CIPN varied depending on the method of CIPN assessment, however older age and higher BMI category were common risk factors across the four models (P<0.05). Age over 60 years was associated with up to 3.3 times increased odds, and overweight/obese BMI status was associated with up to 2.2 times increased odds of chronic CIPN. Diabetes was a risk for patient-reported and neurologically graded CIPN (EORTC-CIPN20, TNSc, NCS, P<0.05) but not clinically graded CIPN (P>0.05). Prior exposure to neurotoxic treatment was a risk factor for CIPN assessed on EORTC-CIPN20, TNSc and NCI-CTCAE, but not NCS (P>0.05). Interestingly, female sex was a significant risk factor for chronic patient reported (β=4.8 95% CI 2.5 – 7.2) and clinically graded CIPN (OR=2.0 95% CI 1.3 – 3.1), whereas male sex was associated with 1.6 times increased odds of reduced sural amplitudes (95% CI 1.0 – 2.4.3, all P<0.05).

**Table 3.** Multivariate models of risk factors associated with chronic CIPN, controlling for time since treatment and chemotherapy type (N=901)

|  | Chronic CIPN Assessment |  |  |  |  |  |  |  |
| --- | --- | --- | --- | --- | --- | --- | --- | --- |
|  | Patient reported measure<br>EORTC-CIPN20 |  | Neurologic evaluation |  |  |  | Clinical grading<br>NCI-CTCAE>0 Y/N |  |
|  |  |  | TNSc |  | Abnormal sural<br>amplitude Y/N |  |  |  |
| Risk Factor | β<br>(95% CI) | P | β<br>(95% CI) | P | Odds ratio<br>(95% CI) | P | Odds ratio<br>(95% CI) | P |
| Age (≥60 vs <60 years) | 4.1 (2.3 – 6.0) | <0.001 | 1.7 (1.3 – 2.1) | <0.001 | 2.0 (1.4 – 2.8) | <0.001 | 3.3 (2.4 – 4.7) | <0.001 |
| Diabetes (Yes vs No) | 3.3 (0.1 – 6.5) | <0.05 | 1.5 (0.8 – 2.2) | <0.001 | 2.8 (1.6 – 5.0) | <0.001 | - | - |
| BMI (compared to<br>underweight/normal) |  |  |  |  |  |  |  |  |
| Overweight | 3.0 (0.9 – 5.1) | <0.01 | 0.5 (0.0 – 1.0) | <0.05 | 1.1 (0.8 – 1.7) | >0.05 | 2.2 (1.5 – 3.1) | <0.001 |
| Obese | 4.0 (1.6 – 6.4) | <0.005 | 1.0 (0.5 – 1.5) | <0.001 | 1.8 (1.2 – 2.8) | <0.01 | 1.8 (1.2 – 2.6) | <0.01 |
| Prior neurotoxic treatment<br>(Yes vs No) | 5.6 (1.8 – 9.3) | <0.005 | 1.4 (0.6 – 2.2) | <0.005 | - | - | 3.2 (1.3 – 7.4) | <0.01 |
| Gender (female vs male) | 4.8 (2.5 – 7.2) | <0.001 | - | - | 0.6 (0.4 – 1.0) | <0.05 | 2.0 (1.3 – 3.1) | <0.005 |
| Pre-existing neuropathy<br>(Yes vs No) | - | - | - | - | - | - | - | - |

### Risk factors for severe CIPN

Risk factors for severe CIPN were investigated by examining participants in the highest quartile of patient-reported CIPN (EORTC-CIPN20; mean score=33.3±21.9), highest quartile of neurologically graded CIPN (TNSc; mean score=9.0±2.0) and moderate/severe clinically graded CIPN (NCI-CTCAE ≥2).

Individual risk factors of severe CIPN are presented in Supplementary Table 4. Older age (≥60 years), higher BMI (overweight and obese compared to underweight/normal) and prior exposure to neurotoxic treatment were associated with significantly increased risk of severe CIPN across all neuropathy measures (all P<0.05).

Multivariate regression models for severe CIPN were investigated, controlling for time since treatment and chemotherapy type (Table 4). Risk factors for developing severe patient reported and clinically graded CIPN included ≥60 years of age (up to 1.8 times odds), higher BMI (up to 2.2 times odds), prior exposure to neurotoxic treatment (up to 2.1 times odds) (all P<0.05). Female sex was only a risk for severe clinically graded CIPN (1.6 times odds, P<0.05). Risk factors for neurologically evaluated severe CIPN were older age (3.1 times odds) and diabetes diagnosis (2.2 times odds) (both P<0.05).

**Table 4.** Multivariate models of risk factors associated with severe chronic CIPN, controlling for time since treatment and chemotherapy type.

|  | **Severe Chronic CIPN Assessment** |  |  |  |  |  |
| --- | --- | --- | --- | --- | --- | --- |
|  | **Highest quartile of patient report (N=224)**<br>**EORTC-CIPN20** |  | **Highest quartile neurological evaluation (N=188)**<br>**TNSc** |  | **Clinically graded mod/severe (N=337)**<br>**NCI-CTCAE $\geq 2$** |  |
| **Risk Factor** | Odds ratio<br>(95% CI) | P | Odds ratio<br>(95% CI) | P | Odds ratio<br>(95% CI) | P |
| Age ($\geq 60$ vs $< 60$ years) | 1.8 (1.3 – 2.5) | $<0.001$ | 3.1 (2.2 – 4.5) | $<0.001$ | 1.8 (1.4 – 2.4) | $<0.001$ |
| Diabetes (Yes vs No) | - | - | 2.2 (1.4 – 3.7) | $<0.005$ | - | - |
| BMI (compared to<br>underweight/normal) |  |  |  |  |  |  |
| Overweight | 1.6 (1.1 – 2.4) | $<0.05$ | - | - | 1.6 (1.1 – 2.2) | $<0.01$ |
| Obese | 2.2 (1.4 – 3.3) | $<0.001$ | - | - | 1.7 (1.2 – 2.5) | $<0.005$ |
| Prior neurotoxic treatment (Yes<br>vs No) | 1.8 (1.0 – 3.3) | $<0.05$ | - | - | 2.1 (1.2 – 3.6) | $<0.05$ |
| Gender (female vs male) | - | - | - | - | 1.6 (1.1 – 2.3) | $<0.05$ |
| Pre-existing neuropathy (Yes vs<br>No) | - | - | - | - | - | - |

## Discussion

Neurotoxicity is recognized as a chronic adverse effect of cancer treatment, and strategies to prevent long-term disability and disruption to QoL are urgently needed. This study investigated clinical risk factors associated with development of dose-limiting, chronic and severe CIPN using a comprehensive multi-modal battery of neuropathy measures. At 12 months post-treatment, our study identified that 70% of patients presented with chronic CIPN, and 37% with moderate-severe symptoms. Prior studies have similarly identified a significant proportion of patients with chronic CIPN (6, 24), representing a large cohort of cancer survivors living with debilitating symptoms. Platinum treated patients experienced more chronic CIPN compared to those treated with taxane or hematological cancer therapies, and CIPN severity was associated with time since end of treatment completion.

### Risk factors for CIPN

Multivariate analyses demonstrated that older age, diabetes diagnosis, higher BMI, and prior exposure to neurotoxic treatment were common risk factors for chronic CIPN across assessment methods, whereas older age, higher BMI and female sex were identified as risk factors for severe CIPN. Similarly, older age and female sex were identified as risk factors for development of dose-limiting CIPN (Figure 4). Patients with moderate-severe clinically graded CIPN also had worse outcomes on patient reported and neurological assessments compared to patients with mild CIPN. Prior studies have also demonstrated that those with severe CIPN report poorer QoL, greater disability and higher falls risk (5, 25). Therefore, while it is important to identify patients who will develop chronic symptoms, there is greater urgency to identify to patients who will develop severe and disabling long-term CIPN.

**Figure 4.**
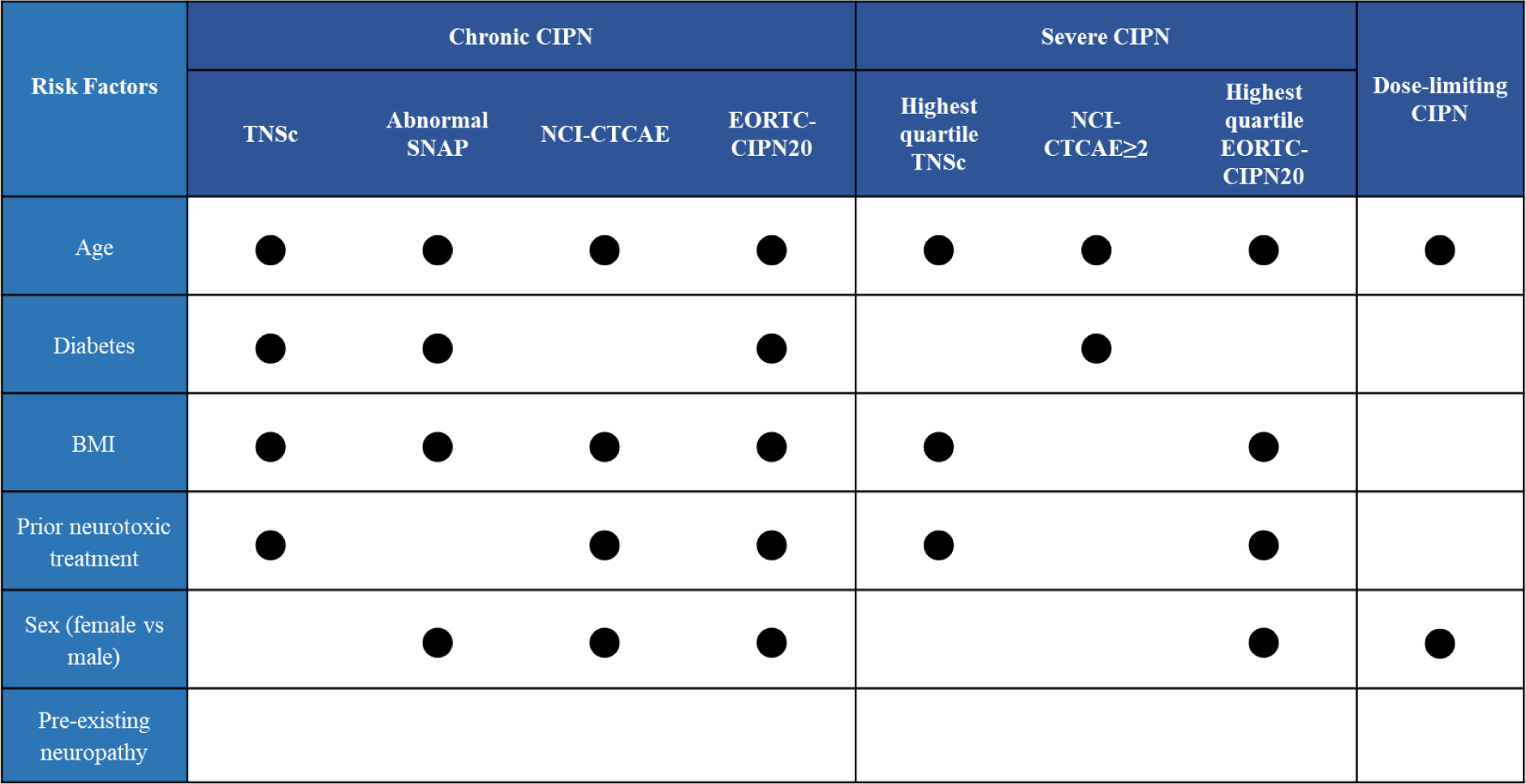
Summary of identified risk factors associated with chronic, severe and dose-limiting CIPN, evaluated using various CIPN assessment approaches.

The profile of patient more likely to develop chronic CIPN differed compared to those likely to develop severe chronic CIPN. While older age and higher BMI were associated with risk of chronic CIPN of any severity, diabetes and prior exposure to neurotoxic therapy were associated with a higher risk of chronic but not severe CIPN. While these risk factors are not modifiable on an individual level, these findings provide clinicians with insights on vulnerable cohorts for closer monitoring, timely intervention and referral to supportive care and neuropathy management (26).

Older age was consistently identified as a risk factor for CIPN – both during and post treatment. Age was associated with higher risk of the development of dose-limiting CIPN, with older people have a 1.4x fold higher risk of dose modification. Age was also associated with the persistence of CIPN post-treatment and associated with severe chronic CIPN. Prior studies have also identified older age as a risk factor for CIPN (12, 13, 27–29). However, older people may have more comorbidities, which may influence risk. Some studies have not identified higher CIPN risk in older people without additional comorbidities (7, 27).

Interestingly, a large-scale study in early breast cancer patients treated with paclitaxel did not confirm any independent association between CIPN and age (30), after controlling for obesity and paclitaxel regimen. Age has been associated with elevated baseline levels of neurofilament-light chain (NfL), a serum-based biomarker of CIPN (31, 32). However future studies will need to investigate whether this increases the risk of NfL elevation during chemotherapy treatment.

Metabolic risk factors have also been associated with CIPN risk (33). This study also identified increased BMI and diabetes to be factors for chronic and severe CIPN. Prior studies have (30, 34, 35) highlighted increased BMI to be associated with an increased 2-3 times risk of CIPN incidence (35) and 1.5 times risk of severe CIPN (30), comparable to results of this study (up to 2.2 times risk for chronic and severe CIPN). Further evidence has also suggested higher BMI increases the burden of CIPN symptoms (36, 37). However not all studies have replicated these results, likely due to the diverse range of CIPN outcome measures used in risk factor analyses (33). Similarly, a prior meta-analysis has demonstrated increased risk (OR=1.6) associated with diabetic status and CIPN (38), however this risk was only significant for paclitaxel (OR=1.5, P<0.05) and not oxaliplatin (OR=1.2, P>0.05) treated patients. The authors suggest this may be due to similarities in pathways of nerve damage between paclitaxel and diabetic peripheral neuropathy (38), although this remains to confirmed. In the present study, diabetes remained a significant risk factor for CIPN after controlling for chemotherapy type (OR=2.2-2.8). Diabetic patients with complications have been demonstrated to be at even greater risk of developing CIPN (OR=2.1) (27). However it is also possible that overall metabolic status, rather than explicitly diabetic and BMI status, influences CIPN risk with studies also highlighting increased larger waist circumference (11) and elevated serum lipids as potential risk factors (39).

Prior treatment with neurotoxic chemotherapies was also identified as a risk factor for CIPN. This has been identified in a prior study (40) where patients had an increased 3.9x risk of CIPN. This is a particular concern for patients with multiple myeloma where neurotoxic regimens are a mainstay treatment for relapsed and refractory disease (41). This study identified up to 3.2x risk of developing chronic CIPN in patients exposed to prior neurotoxic treatment, representing one of the largest risk factor OR identified in this study.

### Mechanisms underlying CIPN risk

The mechanisms by which these factors influence CIPN susceptibility remain unclear. Prior studies have identified associations between older age and higher BMI with increased risk of developing idiopathic neuropathy in the general population (42, 43), suggesting that these factors may be intrinsically associated with neuropathy risk. Aging may specifically increase the risk of neuropathy development following neurotoxic exposure, with studies linking molecular markers of aging to CIPN prevalence (44, 45). Similarly, metabolic factors including hyperlipidemia, hypercholesterolemia and hyperglycemia have also been associated with increased neuropathy risk (46), suggesting that the association between obesity and neuropathy may be mediated by metabolic factors (33). In addition, older or obese patients may be more susceptible to functional disability and less likely to adapt to limitations imposed by CIPN, so that the impact of chronic CIPN symptoms may be higher, influencing perception of risk (11, 47).

This study also identified inconsistent relationship between sex and CIPN risk, depending on how CIPN was assessed. Females were more likely to experience both dose-limiting CIPN and chronic patient reported or clinically graded CIPN. In previous analyses, female cancer survivors reported higher overall symptom burden compared to males (48). Females were also at higher risk of developing painful diabetic neuropathy compared to males (49). These differences may reflect differences in symptom experience or communication. In addition, the study cohort was predominantly female (66%), which may affect findings. Conversely, males were more likely to demonstrate reduced sural amplitudes compared to females. However, males typically have lower SNAP amplitudes than females (50), and may be more likely to reach the cut-of values of the normative range following neurotoxic chemotherapy.

Importantly, polygenic risk factors are likely important in determining CIPN risk, although precise pharmacogenetic pathways have yet to elucidated (51). Genetic contribution to CIPN risk may be agent-specific, due to the different pathophysiological mechanisms associated with neurotoxicity development (52). Multiple single-nucleotide polymorphisms (SNPs) have been associated with CIPN, although meta-analysis failed to find consistent associations, likely related to use of different outcome measures (53). Further studies are required to examine the interplay between metabolic, demographic and genetic and this together impacts on CIPN risk.

### Risk factors for dose-limiting CIPN

The present study also investigated risks associated with dose-limiting CIPN, utilising neurotoxic dose modification due to CIPN as a surrogate marker of significant CIPN during treatment. In line with prior studies (6, 8, 54), 30% of patients in this cohort received dose modification due to CIPN. This represents a significant number of patients whose prescribed anticancer therapy was critically reduced due to CIPN. Identification of this susceptible cohort is an important step to establishing neuropathy management strategies to prolong their treatment exposure. This analysis identified older age and female sex to be associated with increased risk of dose-limiting CIPN, independent of chemotherapy regimen. However, current assessment of CIPN in clinical practice is suboptimal (55), impacting on the reliability of which dose modification decisions are made in line with CIPN status. In fact, studies have demonstrated the benefits of harnessing patient report in clinical in better representing patient’s health status and improving cancer management (56), including increasing duration on chemotherapy (57). It is therefore important to consider incorporating other methods of CIPN assessment into clinical practice, especially given its consequential impact on patients’ exposure to anticancer treatments.

### CIPN assessment approaches

There is currently no clear profile of the patient most likely to develop persisting CIPN, and this may be due to range of CIPN assessment methods utilized across risk factor studies.

While a number of studies used the NCI-CTCAE to grade CIPN (8, 9, 28, 29, 34, 40), patient reported outcome measures, such as FACT/GOG-Ntx (11, 12, 25, 30) as well as the TNSc (11, 12) have also been used to assess neurotoxicity in risk factor analyses. While there is no consensus on the optimal method of CIPN assessment, the NCI-CTCAE is the most commonly used measure in clinical practice and large-scale clinical trials. Although this scale incorporates clinician expertise in benchmarking neuropathy severity (58), the NCI-CTCAE is generally recognised to be a suboptimal measure, lacking sensitivity to change and underreporting symptom severity (55, 59). Training researchers to ensure consistency of CIPN grading may mitigate some pitfalls associated with this measure (60), and this has been adopted in the current study. Furthermore, discordance between CIPN outcome methods have previously been demonstrated (61, 62), with different clinical risk factors identified amongst the same cohort between clinician and patient graded CIPN (63).

This large-scale study adopted a multi-modal CIPN assessment approach incorporating patient reported, neurological and clinical grading of CIPN. Risk factors identified on each assessment approach were not identical, demonstrating the differing constructs of CIPN captured by each assessment. This is further demonstrated in the final multivariate models for severe CIPN, where risk factors for severe patient reported and clinically graded symptoms were the same (older age, higher BMI, prior neurotoxic treatment, female sex), and different to those with severe neurologic CIPN signs and symptoms (older age, diabetes diagnosis).

Given the lack of agreement between these outcome measures, these results are not surprising. These results contribute to the growing evidence suggesting CIPN outcome measures may be assessing differing aspects of neuropathy and emphasises the importance of CIPN assessment selection (16). In the present study, adopting a range of CIPN assessments provides further confidence for the risk factors (i.e. older age and increased BMI) that were identified across all CIPN assessment models.

### Limitations

This study acknowledges a number of limitations. This study recruited patients who had completed their neurotoxic treatment up to five years prior. While this allowed us to capture the impact of chronic CIPN, it is possible that some patients may have developed neuropathy syndromes post treatment. Furthermore, the inclusion of patients with comorbidities including diabetes may have resulted in a heterogenous cohort. However, this study aimed to investigate CIPN risk factors in a real world cancer patient population. This study also recruited from a range of cancer and chemotherapy types, which limits the ability to investigate the effect of chemotherapy-specific factors such as dose as risk factors. Finally, while the present analysis aimed to examine clinical risk factors routinely available at the commencement of cancer therapy, this study acknowledges that clinical risk factors alone may not be able to build a robust CIPN risk model. Other factors including genetic risk factors (51, 53) likely also contribute to prognostic CIPN risk models. Future studies investigating a range of risk types utilising a robust battery of CIPN assessments as in this study will be able to build a more complete model of CIPN.

## Conclusion

The present study has identified baseline clinical risk factors associated chronic, severe and dose-limiting CIPN. These risk factors are routinely available in clinical settings, without the need for specialised assessments or additional resources. Closer monitoring of these vulnerable cohorts will allow for timely CIPN management, including referral pathways to intervention and rehabilitation therapies which will ultimately lead to improved CIPN morbidity.

## Data Availability

All data produced in the present study are available upon reasonable request to the authors

