## Supplementary files for "Peripheral Neuropathy in Cancer Patients- Multifactorial Contributors to Dose Limiting and Chronic Toxicity"

Supplementary Table 1.

Association between time since end of neurotoxic treatment and CIPN outcome measures

| **Outcome measure** | **Spearman correlation** | **Significance** |
| --- | --- | --- |
| EORTC-CIPN20 | -0.1 | <0.005 |
| TNSc | -0.1 | <0.05 |
| NCI-CTCAE grade | -0.1 | <0.005 |

Supplementary Table 2.

CIPN outcome measure scores according by each chemotherapy category

Comparison between chemotherapy categories completed by analysis of variance

Scores presented as mean (SD)

|  | Taxanes  N=518 | Platinums  N=285 | Haematological cancer therapies  N=98 | P |
| --- | --- | --- | --- | --- |
| EORTC-CIPN20 | 13.2 (14.3) | 16.4 (13.8) | 12.1 (10.8) | <0.005 |
| TNSc | 4.1 (3.1) | 4.9 (3.2) | 4.0 (2.9) | <0.005 |
| NCI | 1.1 (0.9) | 1.3 (0.9) | 1.1 (0.9) | <0.005 |
| Abnormal Sural (%) | 23.5% | 52.5% | 43.8% | <0.001 (χ²) |

Supplementary Table 3

Individual clinical risk factors for chronic CIPN, controlling for time since neurotoxic treatment.

*Denotes significant relationship

|  | **EORTC-CIPN20** | | **TNSc** | | **Abnormal Sural (Y/N)** | | **NCI-CTCAE>0 (Y/N)** | |
| --- | --- | --- | --- | --- | --- | --- | --- | --- |
| **Risk Factor** | β  (95% CI) | P | β  (95% CI) | P | Odds ratio  (95% CI) | P | Odds ratio  (95% CI) | P |
| Age (≥60 vs <60 years | 4.2 (2.4 – 6.0) | <0.001* | 1.9 (1.5 – 2.3) | <0.001* | 2.1 (1.5 – 2.9) | <0.001* | 3.0 (2.2 – 4.2) | <0.001* |
| Diabetes | 4.9 (1.7 – 8.0) | <0.005* | 2.2 (1.5 – 2.9) | <0.001* | 3.9 (2.4 – 6.6) | <0.001* | 2.1 (1.1 – 3.9) | <0.05* |
| BMI (compared to underweight/normal) |  |  |  |  |  |  |  |  |
| Overweight | 3.2 (1.1 – 5.4) | <0.005* | 0.7 (0.2 – 1.2) | <0.01* | 1.5 (1.0 – 2.1) | <0.05* | 2.1 (1.5 – 3.0) | <0.001* |
| Obese | 4.7 (2.3 – 7.2) | <0.001* | 1.4 (0.8 – 1.9) | <0.001* | 1.9 (1.3 – 2.9) | <0.005* | 1.8 (1.2 – 2.7) | <0.005* |
| Prior neurotoxic treatment | 5.6 (1.8 – 9.3) | <0.005* | 1.5 (0.7 – 2.3) | <0.001* | 1.8 (1.0 – 3.3) | >0.05 | 3.1 (1.4 – 6.9) | <0.01* |
| Sex (female compared to male) | 1.7 (-0.3 – 3.6) | >0.05 | -0.5 (-1.0 – -0.1) | <0.05* | 0.4 (0.3 – 0.5) | <0.001* | 1.1 (0.8 – 1.5) | >0.05 |
| Pre-existing neuropathy | 7.0 (0.3 – 13.6) | <0.05* | 0.3 (-1.2 – 1.9) | >0.05 | 1.9 (0.6 – 6.0) | >0.05 | 2.9 (0.7 – 12.9) | >0.05 |

Supplementary Table 4.

Individual clinical risk factors for severe chronic CIPN, controlling for time since neurotoxic treatment.

|  | **Highest quartile of EORTC-CIPN20** | | **Highest quartile of**  **TNSc** | | **NCI-CTCAE ≥2 (Y/N)** | |
| --- | --- | --- | --- | --- | --- | --- |
| **Risk Factor** | Odds ratio  (95% CI) | P | Odds ratio  (95% CI) | P | Odds ratio  (95% CI) | P |
| Age (≥60 vs <60 years | 1.9 (1.4 – 2.6) | <0.001* | 3.4 (2.4 – 4.9) | <0.001* | 1.8 (1.3 – 2.3) | <0.001* |
| Diabetes | 1.4 (0.9 – 2.3) | >0.05 | 2.9 (1.8 – 4.7) | <0.001* | 1.7 (1.1 – 2.7) | <0.05* |
| BMI (compared to underweight/normal) |  |  |  |  |  |  |
| Overweight | 1.7 (1.1 – 2.4) | <0.01* | 1.2 (0.8 – 1.8) | >0.05 | 1.6 (1.2 – 2.2) | <0.01* |
| Obese | 2.3 (1.6 – 3.5) | <0.001* | 1.8 (1.2 – 2.8) | <0.01* | 1.8 (1.2 – 2.6) | <0.01* |
| Prior neurotoxic treatment | 1.9 (1.1 – 3.4) | <0.05* | 1.9 (1.1 – 3.5) | <0.05* | 2.2 (1.3 – 3.8) | <0.01* |
| Sex (female compared to male) | 1.2 (0.9 – 1.7) | >0.05 | 0.7 (0.5 – 1.0) | <0.05* | 1.2 (0.9 – 1.6) | >0.05 |
| Pre-existing neuropathy | 2.8 (1.1 – 7.4) | <0.05* | 1.2 (0.4 – 3.8) | >0.05 | 1.5 (0.6 – 4.0) | >0.05 |
